# Stability-related foot placement control relies more on mediolateral center-of-mass velocity feedback in people with early Multiple Sclerosis

**DOI:** 10.64898/2026.08.26.26361464

**Authors:** A.M. van Leeuwen, R. Romijnders, J. Welzel, I. D’Ascanio, K. Stürner, C. Hansen, W. Maetzler

**Affiliations:** Department of Neurology, University Hospital Schleswig-Holstein and Kiel University, Kiel, Germany; Department of Electrical, Electronic, and Information Engineering “Guglielmo Marconi”, University of Bologna, Bologna, Italy

## Abstract

Impaired gait performance and stability is a key symptom often defining disease outcome in people with Multiple Sclerosis. Step-by-step foot placement control in response to variations in the center-of-mass kinematic state is a crucial gait stability mechanism, especially in the mediolateral direction. Even though it is known that people with Multiple Sclerosis are at an increased risk of falling, step-by-step foot placement control remains to be characterized in this population. Here, we explored characteristic foot placement control in ten people with early stage Multiple Sclerosis, compared to 21 controls walking at a similar average gait speed, during 1-minute steady-state treadmill walking. Kinematic data were analyzed using a linear feedback model that correlated foot placement with the center-of-mass kinematic state during the preceding swing phase. People with Multiple Sclerosis demonstrated step-by-step foot placement control in both the mediolateral and anteroposterior directions. No differences were found in foot placement precision between groups. However, foot placement responses to variations in center-of-mass velocity proved stronger in people with Multiple Sclerosis. Moreover, the contribution of mediolateral center-of-mass velocity feedback to the control mechanism was higher in people with Multiple Sclerosis as compared to neurologically healthy controls. Our results suggest that foot placement control is still retained in early clinically evident stages of Multiple Sclerosis, but is realized through differently weighted sensory feedback control.

## Introduction

Multiple Sclerosis (MS) is a chronic immune-mediated disease of the central nervous system, characterized by inflammatory demyelination and progressive axonal and neuronal degeneration disseminated across the central nervous system (1). It is the most common non-traumatic cause of neurological disability in young adults, with typical onset between 20 and 40 years of age and a higher prevalence in women (2, 3). The clinical presentation is highly heterogeneous, spanning sensory, visual, cognitive and motor domains (2, 4). The disease course ranges from relapsing-remitting to progressive forms (3). Among these manifestations, impaired mobility is one of the most frequent and disabling: the majority of people with MS (PwMS) develop gait and balance disturbances over the course of the disease (3, 4), reflecting a combination of muscle weakness, spasticity, impaired proprioception, delayed sensorimotor conduction, impaired sensorimotor integration and cerebellar involvement (4, 5). As a consequence, they are at a high risk of falling (1). Over fifty percent of PwMS fall at least once every year (6), often leading to impacting injuries (6, 7).

During walking, coordinating where we place our foot with respect to the center-of-mass (CoM) is widely considered the most important mechanism for maintaining stability (8, 9). In the anteroposterior direction stable coordination emerges largely from passive dynamics (10, 11), whereas in the mediolateral direction more stabilizing neural control is required (10). Step-by-step foot placement control allows for adjustments in step width and step length in response to swing phase variations in the CoM’s position and velocity with respect to the single stance leg (i.e. the CoM kinematic state) (12). Even in neurologically healthy (young) individuals, such step-by-step control attenuates natural CoM state variation during walking (13). Sensory integration of CoM kinematic state information (14–16) and coordinated muscle activation during the swing phase (13, 17) contribute to precise step-by-step foot placement control.

Neurological impairments in fall prone populations, such as older adults (14), people with Parkinson’s disease (18) and stroke patients (19) have demonstrated detrimental effects on such foot placement control. Similarly, neurological impairments in PwMS may affect step-by-step foot placement control, but has, to our best knowledge, not yet been investigated.

Given the neurological impairments that characterize MS, we hypothesized that in PwMS:

1) Foot placement is coordinated with respect to the center-of-mass kinematic state (H1),

yet, compared to age-matched controls such foot placement control will:

2) be less precise (i.e. a larger magnitude of foot placement errors) (H2).

In addition to testing these hypotheses, we explored the strength of the foot placement responses as well as the contribution of respectively CoM position and CoM velocity information to foot placement control in PwMS.

## Methods

### Participants

Treadmill walking trials, with at least 30 strides (to be able to reliably compute foot placement control outcome measures (20)), from ten people with PwMS and 21 neurologically healthy controls, were included from an existing larger dataset (21). Since foot placement control is speed-dependent (20), controls were selected as such that the mean walking speed was not significantly different, and closely matched between groups (PwMS 1.02 m/s ± 0.3 std; controls: 1.07 m/s ± 0.2 std). On average PwMS scored 1.25 (0 to 4.5; “no disability” to “relatively severe disability”) on the Expanded Disability Status Scale (EDSS) (22). Demographics and clinical data are presented in Table 1.

**Table 1.** Demographics and clinical data.

|  | **People with Multiple Sclerosis** | **Controls** |
| --- | --- | --- |
| *N (Females)* | 10 (4) | 21 (10) |
| *Age [years]* | 35 ± 11 | 30 ± 9 |
| *Height [cm]* | 182 ± 10 | 180 ± 10 |
| *Treadmill walking speed [m/s]* | 1.02 ± 0.3 | 1.07 ± 0.2 |
| *Expanded Disability Status Scale (0-10)* | 1.25 ± 1.4 | N/A |

Data is presented as Mean ± Standard Deviation or Numbers.

As part of an extensive experimental protocol (23), participants walked on two different speeds on the treadmill; 1) comfortable treadmill walking speed (1-minute) and 2) comfortable overground walking speed (1-minute). Comfortable treadmill walking speed was determined by gradually increasing treadmill speed until a speed the participant felt comfortable with (23). Comfortable overground speed was determined prior to the treadmill walking, as the average speed of five 5-meter overground walking trials, given the instruction to walk at their normal walking speed (23).

The included speed for the PwMS was the comfortable overground walking speed (with the exception of comfortable treadmill walking speed for one participant who did not have sufficient strides in the comfortable overground walking speed trial). For the controls we chose the comfortable treadmill (N=14) or the comfortable overground speed (N=7), depending on which speed could be better matched with the PwMS speeds. The first 30 strides of the respective treadmill walking trial were included for each participant.

The ethical board of the medical faculty of Kiel University approved this study (project number D438/18) and all participants gave their written informed consent. The study was registered in the German Clinical Trials Register (DRKS00022998) (21).

### Data collection

Kinematic data was collected using Qualysis at a sampling rate of 200 Hz. Further details can be found elsewhere (23).

### Data preparation

Kinematic data of markers placed on the heels and sternum (21) were used in the analysis. Heel markers were used to determine gait events, based on the treadmill-induced negative velocity of the heel following heel-strike, and the change to positive velocity at toe-off, in accordance with (24). Spline interpolation was applied to recover small gaps (<0.25 s) in the marker data. The sternum marker was used as a proxy for the trunk CoM, as was previously done using thorax markers in (25, 26).

### Foot placement models

Step-by-step foot placement control can be characterized using the “foot placement model” (12), a linear regression correlating CoM kinematic state (i.e. CoM position and velocity with respect to the stance foot) and subsequent mediolateral (i.e. step width, *SW)* [equation 1] or anteroposterior (i.e. step length, *SL*) foot placement [equation 2].

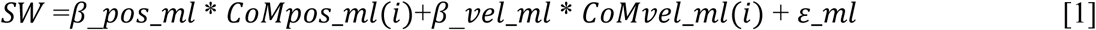

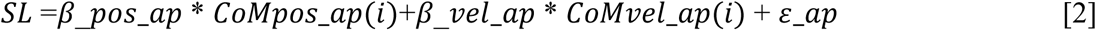

*β*_*pos* and *β*_*vel* denote the the regression coefficients/control gains, of respectively the CoM position (*CoMpos*) and velocity (*CoMvel*). *ε*_*ml* and *ε*_*ap* are the residuals (i.e. “foot placement errors”) for the mediolateral and anteroposterior direction. *i* represents the step percentage for which the model is fitted. Each step is time-normalized to 51 samples, representing 100% of the swing phase (from toe-off to ipsilateral heelstrike). We fitted the model for samples *1*, *25* and *51*. Whereas the model fitted at the end of the step (i=51) generates measures which are generally considered to be related to foot placement control performance (i.e. how well is foot placement ultimately coordinated with respect to the CoM), models fitted at earlier step percentages (e.g. at the start of the step *i=1* and at mid-swing *i=25*) are considered giving insight in the underlying feedback control (i.e. how well does the CoM state during the start of the swing phase and during mid-swing predict the upcoming foot placement.

The standard deviation of the residuals (*ε_ml_*, *ε_ap_*) can be considered as the *magnitudes of foot placement errors*, and as such as measures of foot placement precision (27). The *magnitude of the regression coefficients* of the foot placement model were evaluated as a measure of the strength of the foot placement responses (27). *Partial correlations* of the predictors (CoM_pos,_ CoM_vel)_ provide insight in the overall quality of foot placement control, as well as the relative contributions of position and velocity information to foot placement control (27, 28).

### Statistics

Given the small PwMS sample size, we considered our investigation exploratory and used Bayesian statistics (29), allowing us to report our findings under the available “degree of evidence” given the current sample (29, 30). The degree of evidence is denoted by the Bayes Factor (BF) and ranges from “Anecdotal” to “Extreme”, in accordance with (29). In case of “Anecdotal” evidence the results of this study were considered inconclusive (29). All statistical analyses were performed in JASP (Version 0.18.3) (31), using the default Cauchy prior distribution (centered around 0 with a scale parameter of 0.707). JASP provides Bayesian equivalents of one-sided and two-sided t-tests yielding respectively a BF_+0_/BF_0+_/ BF_-0_/BF_0-_ or BF_10_/BF_01_, representing support for the alternative or the null hypothesis, respectively.

To provide initial context, we compared average step width and length between groups using the Bayesian equivalent of a one-sided independent t-test.

To test H1, whether PwMS use step-by-step foot placement control, we tested the regression coefficients against zero, similar to (17) and (28), using the Bayesian equivalent of a one-sample t-test.

To test H2, that foot placement precision would be diminished in PwMS, we compared the standard deviation of the residuals (*ε_ml_*, *ε_ap_*; i.e. foot placement errors) between groups, using the Bayesian equivalent of a one-sided independent t-test.

To explore the strength of the foot placement responses in PwMS as compared to controls, we compared the regression coefficients between groups, using the Bayesian equivalent of a two-sided independent t-test.

To explore the contributions of respectively CoM position and CoM velocity information to foot placement control, we tested the respective partial correlations between groups, using the Bayesian equivalent of a two-sided independent t-test.

To capture both the feedback control, as well as the resulting foot placement outcome, we performed all tests separately for the models fitted at the start of the step (start_step), at mid-swing (mid_step) (considered as feedback control phase) and at the end of the step (end_step, considered as the foot placement outcome), respectively.

## Results

The results cover the statistically tested step percentages. Illustrations of the outcome measures across the full step cycle can be found in the *Supplementary material*.

**Figure 1.**
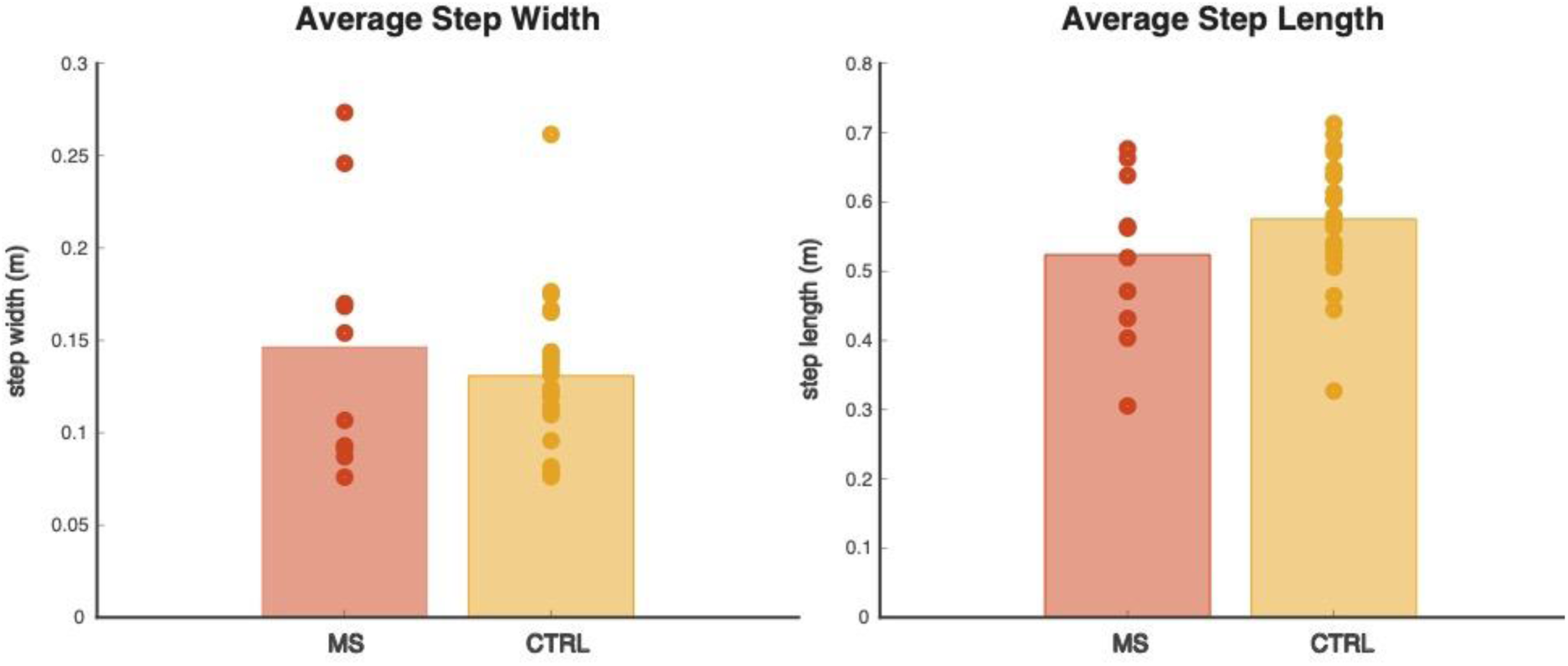
Step width and step length. Step width and step length in meter. People with Multiple Sclerosis (MS, red) and controls (CTRL, yellow) showed similar step width and anecdotal shorter step length. Circles represent individual data points.

### Step width and step length

We found anecdotal evidence (BF_+0_ = 0.7) supporting comparable step width and anecdotal evidence (BF_-0_ = 1.2) supporting shorter step length in PwMS as compared to controls.

### Step-by-step foot placement control

#### CoM_pos_ regression coefficient

In the mediolateral direction, we found extreme evidence for regression coefficients larger than zero at the start of the step (BF_+0_ = 12990), at mid-swing (BF_+0_ = 18413) and at the end of the step (BF_+0_ = 273960), indicative of retained step-by-step foot placement control with respect to the CoM kinematic state in PwMS (H1). In the anteroposterior direction, PwMS also demonstrated step-by-step foot placement control (H1), as evidenced by extreme evidence at the middle and end of the step (BF_+0_ = 2706; BF_+0_ = 1530). However, at the start of the step only anecdotal evidence was found (BF_+0_ = 2.3) supporting a regression coefficient larger than zero, but remaining inconclusive.

Similar results were found for the control group. For the mediolateral direction, extreme evidence supported the execution of the step-by-step foot placement control mechanism, when tested for the start of the step (BF_+0_ = 4.1 *10^6^), at mid-swing (BF_+0_ BF_+0_ = 1.4*10^8^) and at the end of the step (3.5*10^8^). In the anteroposterior direction, they demonstrated step-by-step foot placement control when tested at mid-swing (very strong evidence: BF_+0_ = 88) and the end of the step (extreme evidence: BF_+0_ = 102150), but only anecdotal evidence was found at the start of the step (BF_+0_ =1.7).

**Figure 2.**
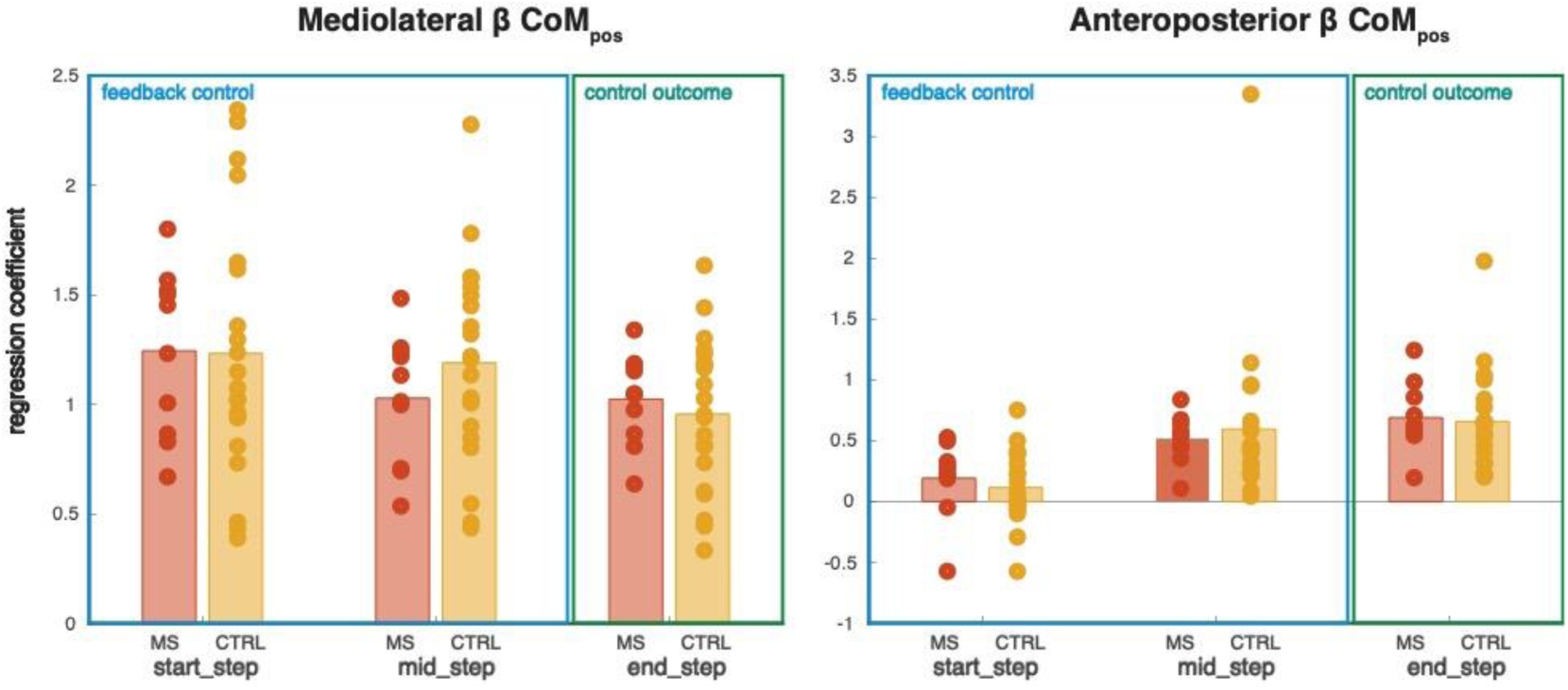
Mediolateral and anteroposterior relationship between CoM position and foot placement. CoM position regression coefficients (β CoM_pos_) for People with Multiple Sclerosis (MS, red) and controls (CTRL, yellow), for the models fitted at start_step, mid_step (interpreted as feedback control:blue) and end_step (interpreted as control outcome:green). Circles represent individual data points.

#### CoM_vel_ regression coefficient

In PwMS, for the mediolateral direction, extreme evidence supported the relationship between CoM velocity and foot placement, at the start of the step (BF_+0_ = 3293), at mid-swing (BF_+0_ = 27329) and at the end of the step (BF_+0_ =2166). In the anteroposterior direction, we found strong evidence at the start of the step (BF_+0_ = 12), anecdotal evidence at mid-swing (BF_+0_ = 1.5) and strong evidence at the end of the step (BF_+0_ = 20).

For the control group, we also found mediolateral foot placement control based on CoM velocity information, given extreme evidence at the start of the step (BF_+0_ = 6613) at mid-swing (BF_+0_ = 5911), and at the end of the step (BF_+0_ =22434). In the anteroposterior direction we found extreme evidence at the start of the step (BF_+0_ = 2180) and extreme evidence at the end of the step (BF_+0_ =30529), for regression coefficients larger than zero. When testing at mid-swing we found anecdotal evidence against a larger regression coefficient than zero (BF_+0_ = 0.8).

Overall, these results support H1; PwMS execute step-by-step foot placement control during steady-state walking.

**Figure 3.**
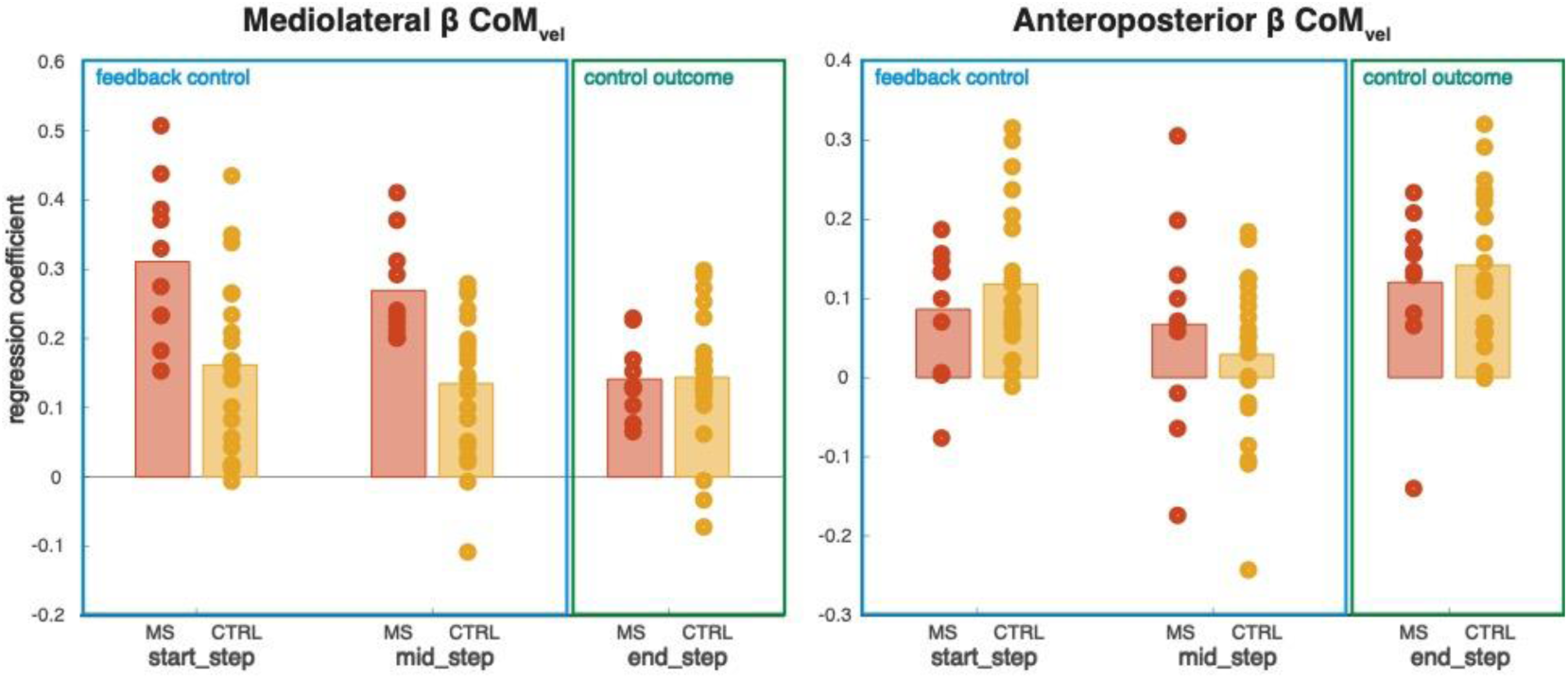
Mediolateral and anteroposterior relationship between CoM velocity and foot placement. CoM velocity regression coefficients (β CoM_vel_) for People with Multiple Sclerosis (MS, red) and controls (CTRL, yellow), for the models fitted at start_step, mid_step (interpreted as feedback control:blue) and end_step (interpreted as control outcome:green). Circles represent individual data points.

### Foot placement precision

At the start of the step, mid-swing and at the end of the step we found anecdotal evidence (start_step, BF_+0_ = 0.6; mid_step, 0.4; end_step, 0.5) supporting similar mediolateral foot placement errors in PwMS as compared to controls (Figure 4, left panel). Similarly, for the anteroposterior foot placement errors anecdotal evidence was found (start_step, BF_+0_ = 0.5; mid_step, 0.6; end_step, 0.7) supporting similar anteroposterior foot placement errors in PwMS as compared to controls (Figure 4, right panel).

Given the anecdotal evidence these results are inconclusive regarding H2, whether foot placement errors are higher in PwMS than in controls or not.

**Figure 4.**
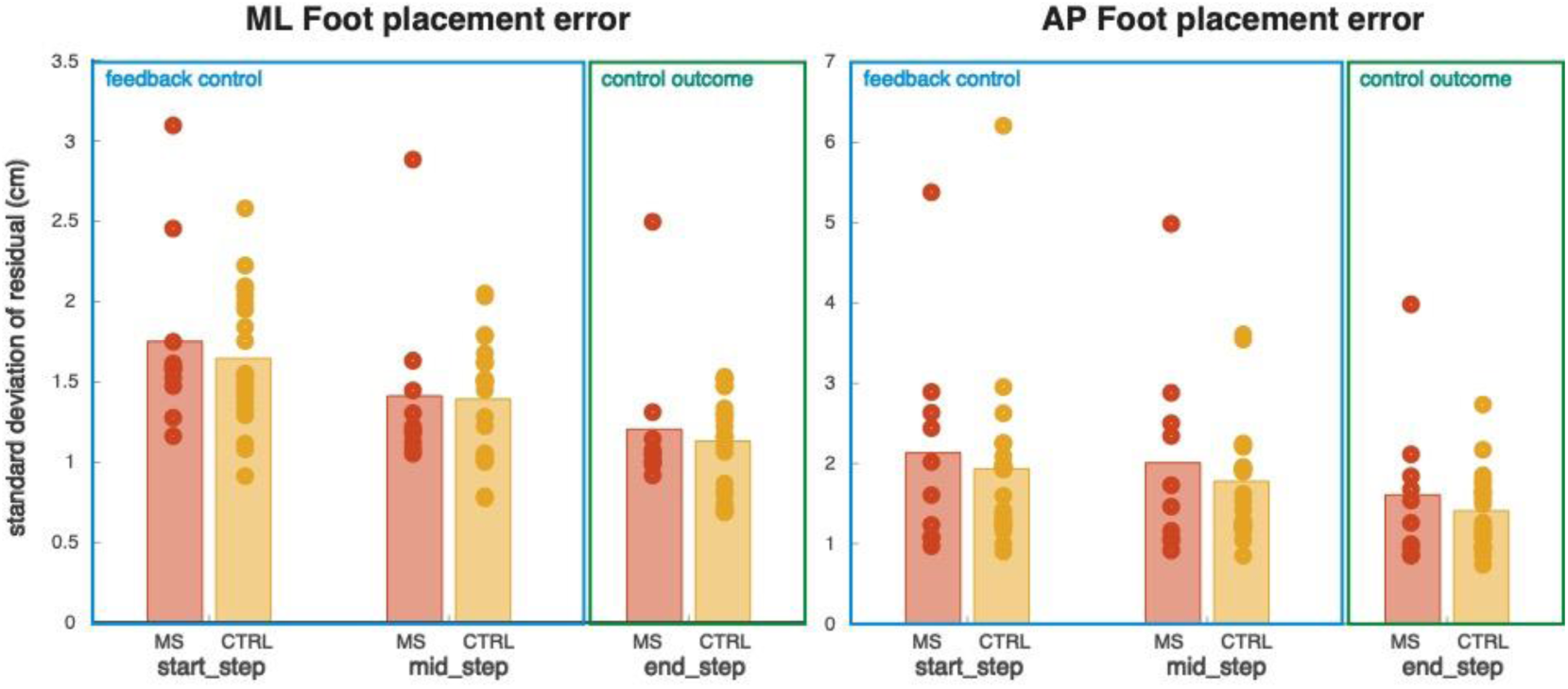
Mediolateral and anteroposterior foot placement precison. Foot placement errors as a measure of foot placement precision for People with Multiple Sclerosis (MS, red) and controls (CTRL, yellow), for the models fitted at start_step, mid_step (interpreted as feedback control:blue) and end_step (interpreted as control outcome:green). Circles represent individual data points.

### Strength of foot placement responses

#### Responses to deviations in CoM_pos_

For mediolateral foot placement control, we found anecdotal evidence at the start of the step (BF_10_ = 0.4), at mid-swing (BF_10_ = 0.5), and at the end of the step (BF_10_ = 0.4), supporting similarly strong responses to deviations in CoM position in PwMS and controls (Figure 2, left panel). Comparable results were found in the anteroposterior direction (Figure 2, right panel), where we found anecdotal evidence supporting similarly strong responses in PwMS and controls, at the start of the step (BF_10_ = 0.4), at mid-swing (BF_10_ =0.4), and at the end of the step (BF_10_ =0.4).

#### Responses to deviations in CoM_vel_

Concerning the response to deviations in CoM velocity in the mediolateral direction, we found strong evidence at the start of the step (BF_10_ = 12) supporting stronger foot placement responses in PwMS than in controls. Similarly, at mid-swing very strong evidence (BF_10_ = 32) supported stronger responses in PwMS to deviations in CoM velocity, than in controls. In contrast, at the end of the step we found anecdotal evidence (BF_10_ = 0.4) supporting similarly strong responses in PwMS and controls (Figure 3, left panel). For the anteroposterior direction, we found anecdotal evidence supporting similarly strong responses for PwMS and controls at the start of the step (BF_10_ = 0.5), at mid-swing (BF_10_ = 0.5) and at the end of the step (BF_10_ = 0.4; Figure 3, right panel).

### Contributions of CoM position and CoM velocity

In the mediolateral direction, the contribution of *CoM*_pos_ tested similar for PwMS and controls, but only based on anecdotal evidence (start_step, BF_10_ = 0.4; mid_step, BF_10_ = 0.4; end_step, BF_10_ = 0.6), deeming this result inconclusive. For the anteroposterior direction, we found similar anecdotal evidence (start_step, BF_10_ = 0.6; mid_step, BF_10_ = 0.4; end_step, BF_10_ = 0.4).

**Figure 5.**
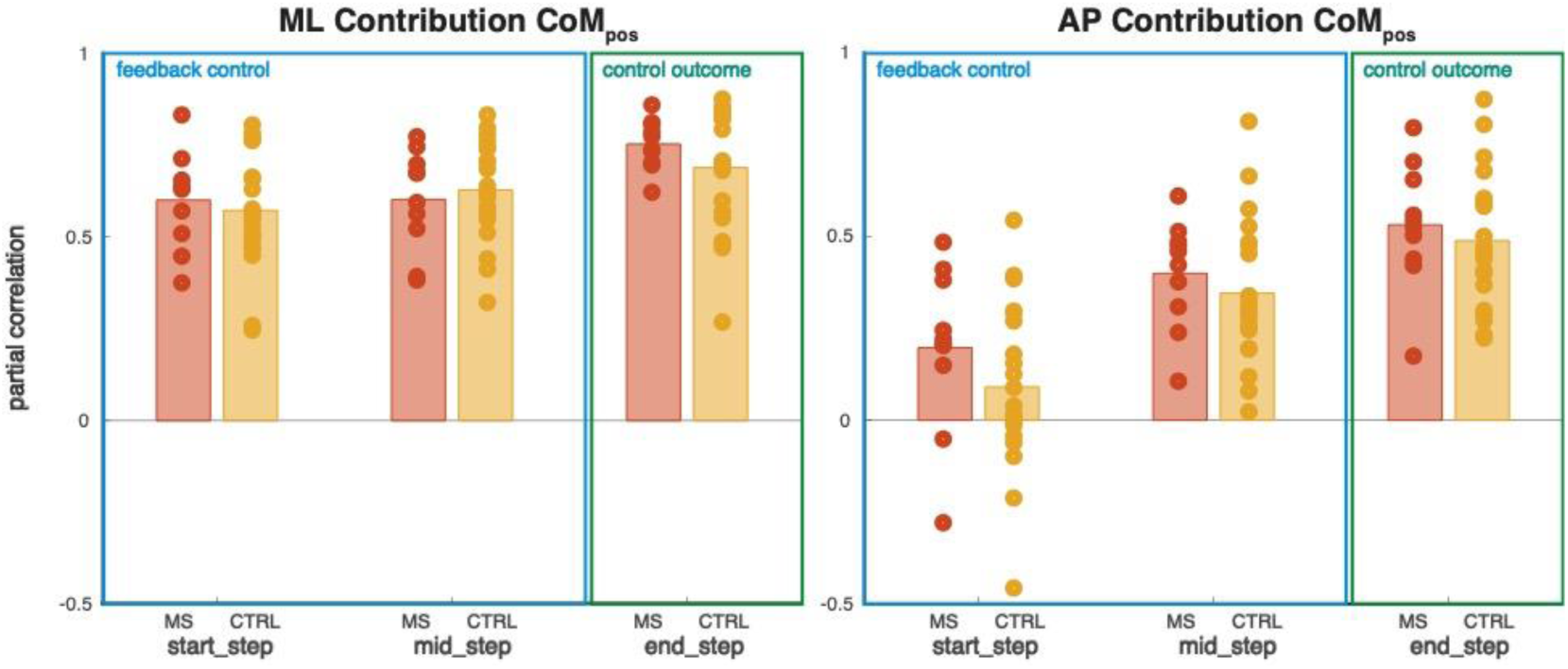
Mediolateral and anteroposterior contribution of CoM_pos_. Partial correlations, as a measure of feedback/control contribution of CoM_pos_. Bars represent the partial correlations of respectively People with Multiple Sclerosis (MS, red) and controls (CTRL, yellow), for the models fitted at start_step, mid_step (interpreted as feedback control:blue) and end_step (interpreted as control outcome:green). Circles represent individual data points.

For the mediolateral direction we found strong evidence supporting a larger contribution of CoM velocity information at the start of the step (BF_10_ = 14) and mid-swing (BF_10_ =16). At the end of the step the results were inconclusive; anecdotal evidence pointed towards similar contributions in PwMS and controls (BF_10_ = 0.4). For the anteroposterior direction, anecdotal evidence pointed towards similar CoM velocity information in PwMs and controls at all three tested gait percentages (start_step, BF_10_ = 0.4; mid_step, BF_10_ = 0.5; end_step, BF_10_ = 0.4). As such, the results for anteroposterior foot placement control remained inconclusive.

**Figure 6.**
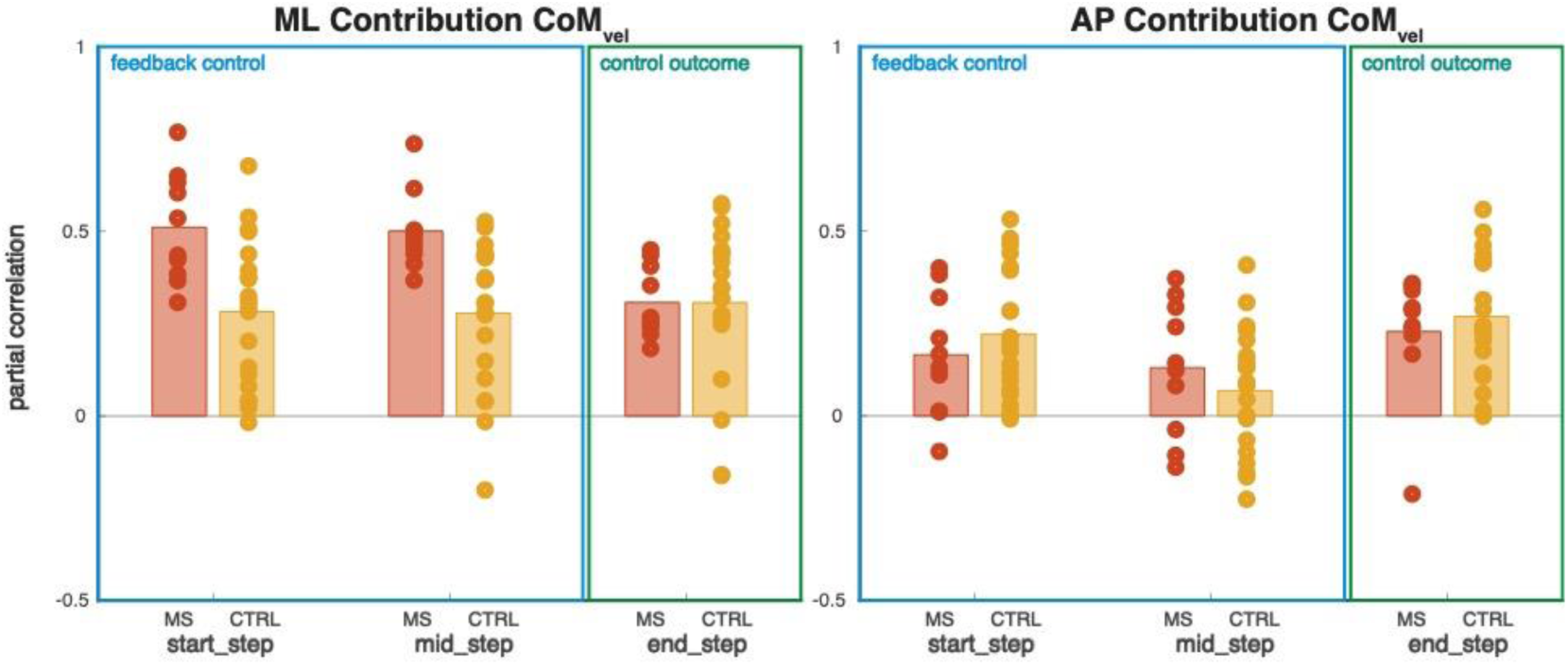
Mediolateral and anteroposterior contribution of CoM_vel_. Partial correlations, as a measure of feedback/control contribution of CoM_vel_. Bars represent the partial correlations of respectively People with Multiple Sclerosis (MS, red) and controls (CTRL, yellow), for the models fitted at start_step, mid_step (interpreted as feedback control:blue) and end_step (interpreted as control outcome:green). Circles represent individual data points.

## Discussion

We were able to demonstrate the preservation of step-by-step foot placement control in this group with early PwMS in both the mediolateral and anteroposterior direction. Our results demonstrate that PwMS rely more on CoM velocity feedback to achieve such control, and respond more strongly to variations in CoM velocity compared to controls.

### Included PwMS

The PwMS included in the current study had a low average EDSS score (mean: 1.25), indicating that they were a relatively well-functioning group of PwMS in an early disease stage. In contrast to what was found in the literature (32), we did not find conclusive evidence that this group of PwMS walked with wider and shorter steps as a compensatory stability mechanism (Figure 1). Apart from low disease severity (32), treadmill walking may have masked earlier reported step width differences between PwMS and controls, as walking on a split-belt treadmill is known to increase step width in controls (33). In contrast, since PwMS already walk with an increased step width during overground walking (32), walking on a split-belt treadmill may not have further increased their step width, making group differences in step width more ambiguous. Moreover, in the current study the groups were matched based on walking speed, whilst slower walking in PwMS may explain the earlier reported shorter step length compared to controls (32). Overall, based on these aspects, we consider the results of this research valid and relevant for understanding early foot placement control adaptation during this disease, which has to be further evaluated concerning clinical implications.

### Foot placement control performance: comparable between PwMS and controls

Step-by-step foot placement control is speed-dependent, with a lower degree of foot placement control at slower speeds (17, 20). By comparing a group of PwMS and a group of controls walking at similar treadmill speeds, we were able to draw conclusions regarding how MS affects foot placement control. In line with H1, PwMS demonstrated step-by-step foot placement control in both the mediolateral and anteroposterior direction (Figure 2). In both PwMS and controls such control was apparent from the start of the step in the mediolateral direction. In the anteroposterior direction conclusive evidence was found only at mid-swing and at the end of the step, suggesting that anteroposterior foot placement control commences later in the gait cycle. A modelling study postulated that both the position and velocity regression coefficients need to be positive to manifest local stability (34). From Figures 2 and 3, it becomes apparent that this is not the case for all participants. This indicates that not all participants prioritized stability constraints when controlling foot placement. It has been reported before that an additional foot placement task diminishes the overall quality of foot placement control (35). For example, in the anteroposterior direction one may prioritize keeping up with the treadmill speed. In such situations, other stability mechanisms can complement foot placement control (36, 37). Given the context of our analysis, some controls may not have prioritized stability related foot placement control, as at a slow speed they might not perceive high stability demands. From the literature however, we know that PwMS prioritize a large margin of stability (32), and this may be reflected here again, as for the mediolateral direction, we found only positive regression coefficients for all PwMS.

We had hypothesized that PwMS would demonstrate less precise foot placement control (H2). Although Figure 4 illustrates a trend towards slightly higher foot placement errors in PwMS, the Bayesian evidence remains inconclusive and even teases at similar foot placement errors between groups. This suggests that, at the speeds as observed in the current study, relatively well-functioning PwMS (mean EDSS = 1.25) are able to control foot placement as precisely as controls. This does not license the conclusion that PwMS and controls have equivalent foot placement capacity in general and under all environmental and personal conditions. Participants walked at relatively slow speeds (32), and neurologically healthy individuals tend to lower their degree of foot placement control at such speeds (20). Moreover, slowing down walking speed in PwMS may be a compensatory strategy, allowing them to operate at a speed for which their foot placement precision suffices. Again, maintaining a large margin of stability seems a priority for PwMS (32) and precise step-by-step foot placement control can contribute to achieving this. Whether PwMS can sustain similar precision to controls when pushed, for example at faster-than-comfortable speeds, should be tested in future work. Moreover, given the association of non-motor aspects, particularly fatigue, in MS with fall risk (4), future work should also assess the influence of such symptoms on foot placement control in MS.

### Foot placement control and sensory feedback: differences between PwMS and controls

Although foot placement control outcomes (Figures 2&3) and precision (Figure 4) presented similarly in PwMS and in controls, there were differences between groups observed in how such precise foot placement outcomes were achieved. For the mediolateral direction, we found that PwMS responded more strongly to variations in CoM velocity at the start of the step and at mid-swing (when feedback control for foot placement happens; Figure 3). Moreover, the contribution of CoM velocity information in predicting mediolateral foot placement was larger in PwMS than controls (Figure 6), at the start of the step and at mid-swing (again when feedback happens), but not at the end of the step (when the “result” of the foot placement control occurs). Finding such differences throughout the swing phase (start of the step, mid-swing), rather than at the end of the step, suggests a stronger reliance on sensory processing of CoM velocity feedback to control foot placement (38) in PwMS as compared to controls, whilst targeting a similar control outcome as the controls (end of the step).

Information about the CoM kinematic state (i.e. CoM position and velocity) can be inferred through sensory integration of input from different sensory modalities (14–16). Proprioceptive, vestibular and visual inputs have all been shown to influence foot placement control (14–16). In particular, impaired proprioceptive feedback in PwMS (4) may affect foot placement control. Due to the cumulative impact of lesions along the central neural pathways to the lower limb in MS, proprioception is often impaired (5). In addition, spasticity, sometimes even present in mildly affected PwMS (39), can degrade the reliability of position-based proprioceptive feedback through secondary mechanical changes in spastic muscle (such as increased passive stiffness and altered thixotropic properties, (40, 41)). Assuming that proprioceptive feedback is indeed impaired in this cohort, our results would indicate that successful sensory reweighting (42) retained foot placement control. This would explain earlier reported stronger effects of visual flow perturbations during treadmill walking in PwMS (43). Visual flow holds information about CoM velocity during gait (44). In this light, both the perturbation study, and the larger contribution of CoM velocity feedback in the current study may point towards a higher reliance on visual feedback during walking in PwMS.

Alternatively, spasticity may contribute to potential reweighting of CoM velocity information through a different route. Spasticity is velocity-dependent: the exaggerated stretch reflexes that characterize it arise preferentially for fast stretches, reflecting disinhibition of velocity-sensitive Ia pathways through reduced presynaptic and reciprocal inhibition rather than a primary change at the receptor (45, 46). As such, the disinhibition of velocity-sensitive afferent pathways might itself amplify the weighting of velocity-related information.

To better understand the nature of the observed sensory reweighting in PwMS, probing of the vestibular, proprioceptive and visual systems would be the logical next research step. In case of down-weighted proprioceptive feedback, re-training of proprioception may support walking stability in PwMS at a later stage of disease progression. A sensory augmentation intervention tailored to step-by-step foot placement control exists (27, 47), and may as such be beneficial for PwMS. Further neural characterization of foot placement control in PwMS could be achieved through electromyography and EEG recordings during foot placement control (48). Foot placement control in MS is expected to have neural (-timing) signatures we did not measure here. The characteristic early-swing burst of gluteus medius activity associated with mediolateral foot placement (13, 17) may be delayed in PwMS given slowed conduction (5). Moreover, cortical involvement may be greater if maintaining balance draws more heavily on attentional resources, consistent with the tendency of PwMS to fall under fatigue and cognitive load (4).

### Foot placement control and complementary mechanisms: Impaired ankle moment control in PwMS

Apart from directly affecting foot placement related proprioception, spasticity may indirectly lead to sensory upweighting of CoM velocity feedback by impairing ankle moment control in PwMS (49). MS may dominantly hamper step-by-step ankle moment control, which relies on fast, reflex-like adaptations in ankle moments during stance (37, 50). Neurologically healthy adults can rely on ankle moment control to compensate for foot placement errors (37, 51). In PwMS, co-contraction of the tibialis anterior and triceps surae muscles reduces ankle mobility (49), and may therefore prevent effective step-by-step ankle moment control. Ankle moment control has been constrained experimentally before (17), effectively making people less stable (26, 52), and putting a higher stabilizing demand on foot placement control (34). Curiously, such constrained ankle moment control led to stronger foot placement responses in response to variations in CoM velocity (i.e. a larger β_vel_ml_), but not in response to variations in CoM position (i.e. no significant effect on β_pos_ml_) (34). Moreover, walking with wider steps and at a higher step frequency appear to be general compensatory mechanisms when ankle moment control is constrained (17). These compensatory mechanisms resemble what has been earlier reported related to PwMS (i.e. wider and shorter steps) (32), in combination with the main result of the current study (i.e. reweighting of CoM velocity feedback in PwMS). Therefore, a lack of complementary ankle moment control in PwMS appears a likely explanation for the observed results. As such, our study motivates research into other gait stability mechanisms, such as ankle moment control (37, 53), and the investigation of the potential to train such mechanisms in PwMS to reduce fall risk.

### Strengths and limitations

By matching PwMS and controls on treadmill walking speed, we could attribute group differences to disease aspects and effects, rather than to speed, as the latter is known to strongly modulate foot placement control (20). However, there are also limitations. For example, our sample was small. We addressed this insofar as we treated the analysis as exploratory and Bayesian throughout, reporting findings under their available degree of evidence. Yet, some results remained anecdotal and require a larger sample size before strong conclusions can be drawn. Another limitation was the restriction of the study to PwMS who could sustain sufficient strides on a treadmill. As a consequence, the MS group was only minimally functionally impaired (mean EDSS = 1.25) and the findings may not generalise to more severely affected PwMS. Moreover, this being a treadmill study introduces further limitations related to generalizability. Foot placement control manifests itself differently during overground as compared treadmill walking (28). Specifically, the contribution of CoM velocity to the foot placement control mechanism becomes more important during overground walking. It remains thus to be investigated how foot placement control is achieved, and how accurate it is, during overground walking in this cohort. Lastly, throughout this paper we interpret the foot placement models used as “feedback control” models, as in (38). This interpretation has been supported by sensory perturbation studies (14, 15, 43). It must be noted however, that in the current work we are unable to formally distinguish passive (i.e. due to the mechanical coupling between the CoM and the swing foot) and neural contributions (i.e. muscle-driven foot placement adjustments during swing (13, 17)) to foot placement control. Whilst for anteroposterior foot placement control passive dynamics may dominate the correlations (in Figures 2&3) rather than feedback control (11), neural contributions to mediolateral foot placement control have been well-established in the literature (10, 13, 17).

## Conclusion

In this treadmill gait-based study, early stage PwMS retained step-by-step foot placement control as a stabilizing mechanism during steady-state gait. When compared against neurologically healthy persons walking at a similar walking speed, they appear to achieve similar foot placement precision. However, in PwMS a larger contribution of CoM velocity feedback underlies this control. Future research and training interventions should focus on further investigating this phenomenon also in more advanced stages of the disease, including the evaluation of sensory integration and sensory re-weighting strategies.

## Data Availability

The dataset used in the present study is available upon reasonable request to the authors, and after signing a data sharing agreement: https://doi.org/10.3390/data7100136

## Supplementary material

### Foot placement precision

**Supplementary Figure 1.**
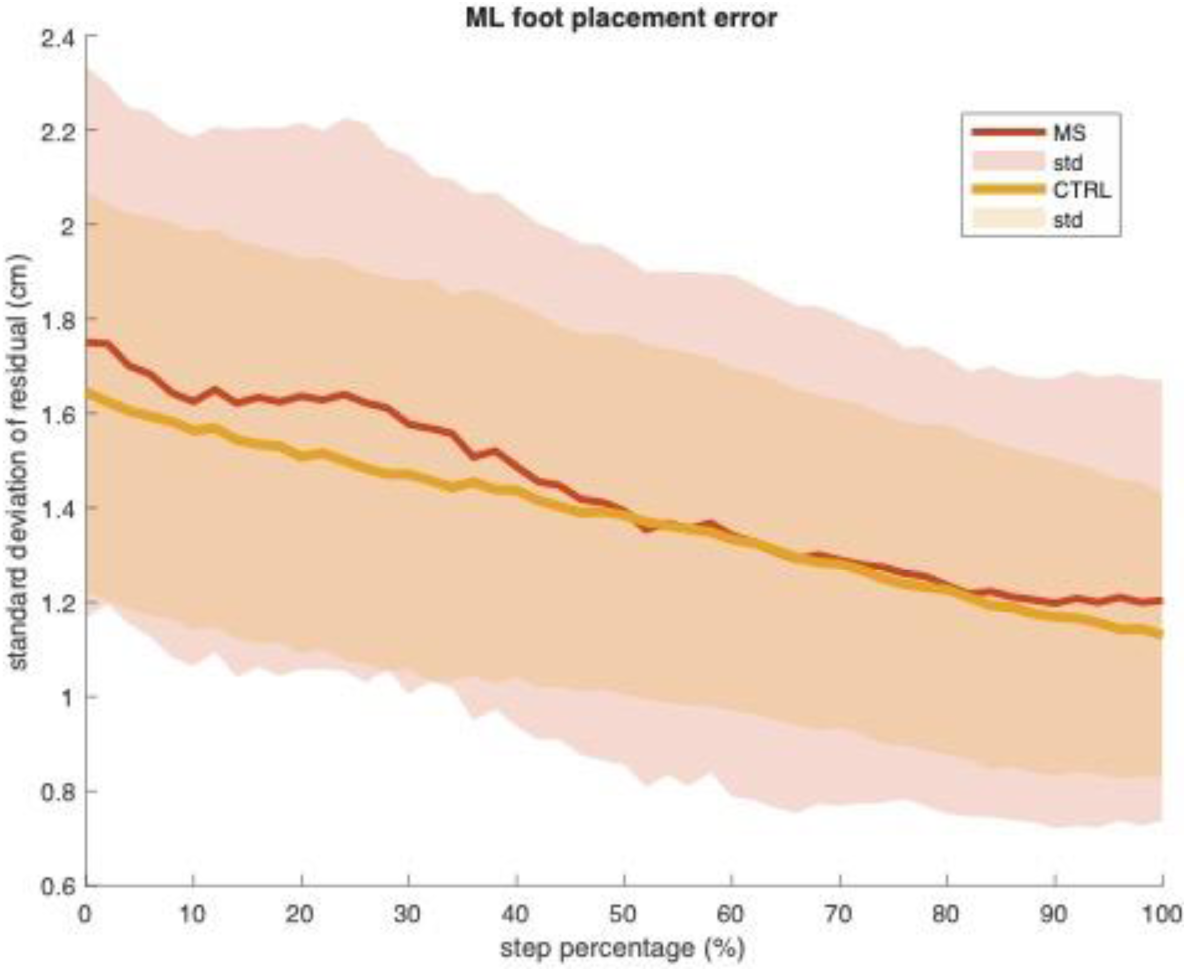
Mediolateral foot placement error. Mediolateral foot placement error, as a measure of foot placement precision, across the step cycle. PwMS (red) and controls (yellow) show similar foot placement error magnitudes.

**Supplementary Figure 2.**
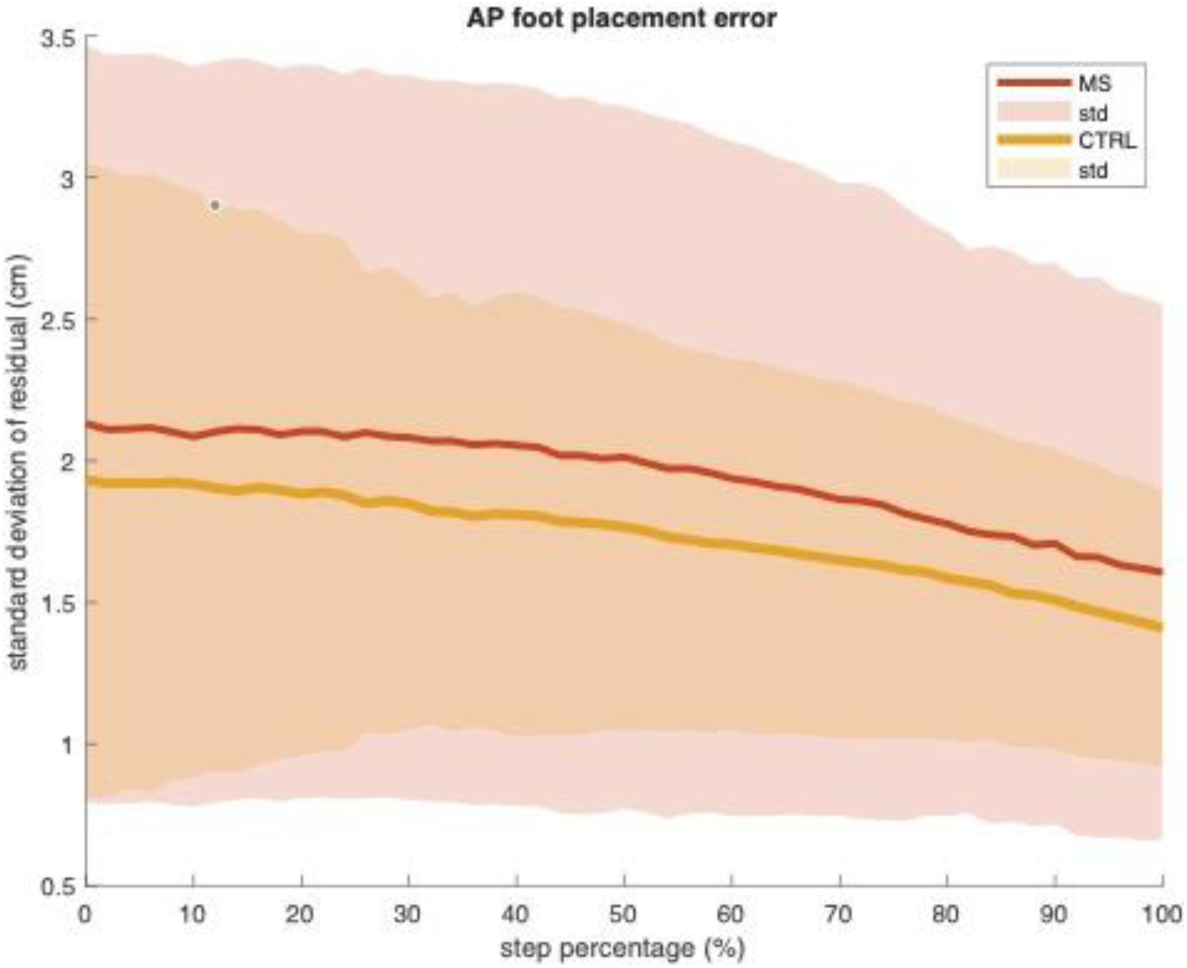
Anteroposterior foot placement error. Mediolateral foot placement error, as a measure of foot placement precision, across the step cycle. PwMS (red) and controls (yellow) show similar foot placement error magnitudes.

### Regression coefficient CoM position

**Supplementary Figure 3.**
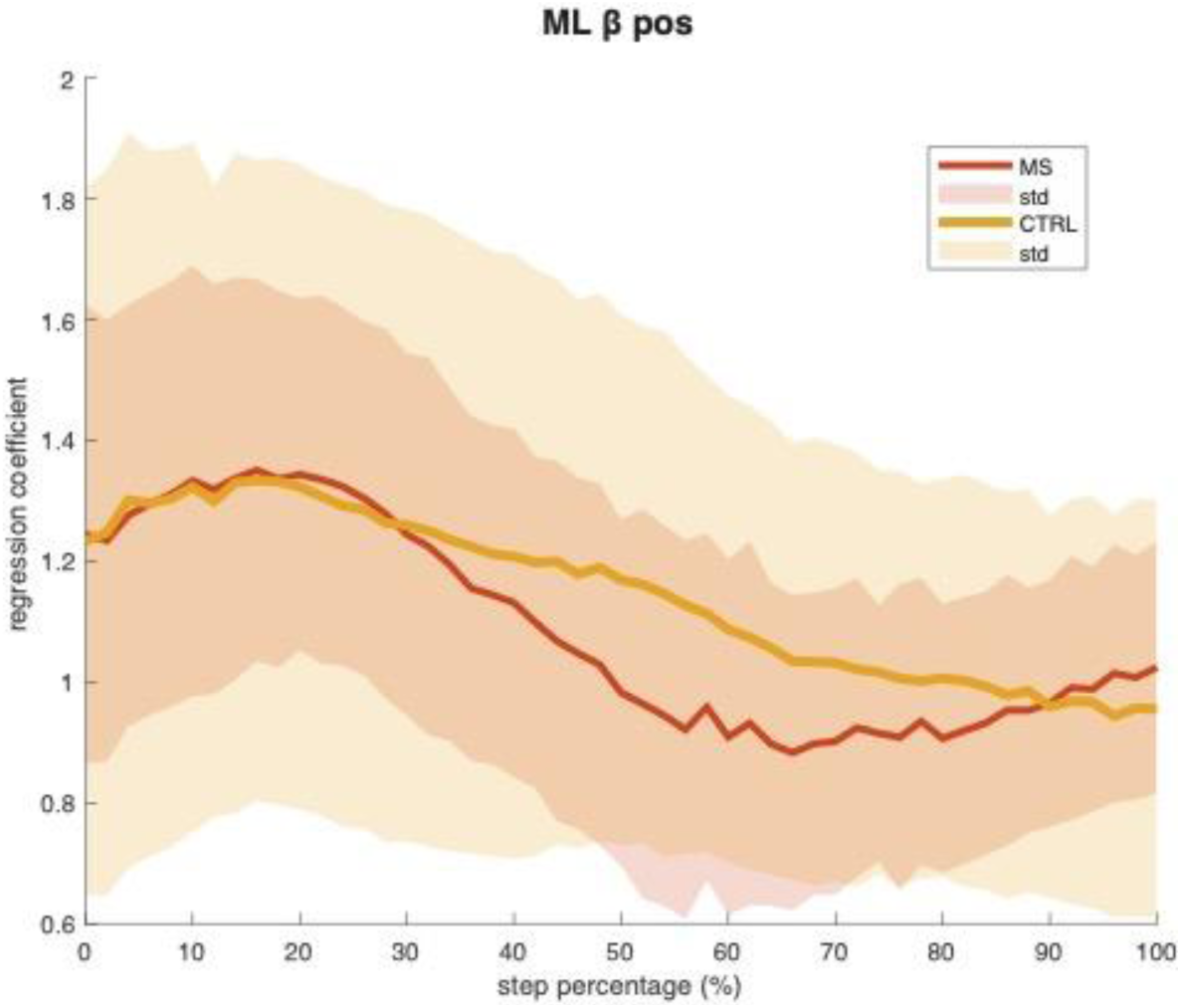
Mediolateral regression coefficient CoM_pos_. CoM_pos_ regression coefficient, as a measure of the strength of foot placement responses, across the step cycle, in PwMS (red) and controls (yellow).

**Supplementary Figure 4.**
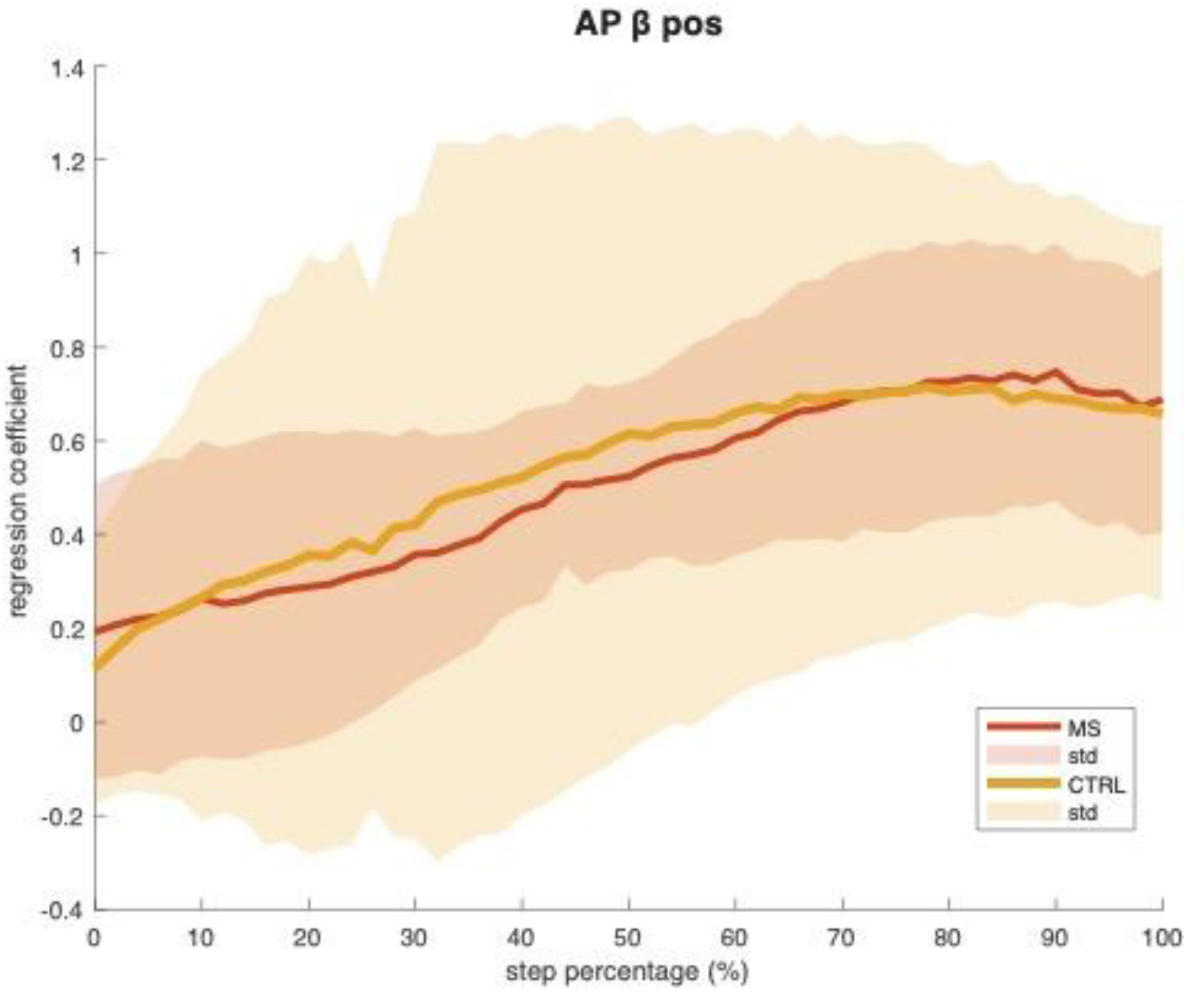
Anteroposterior regression coefficient CoM_pos_. CoM_pos_ regression coefficient, as a measure of the strength of foot placement responses, across the step cycle, in PwMS (red) and controls (yellow).

### Regression coefficient CoM velocity

**Supplementary Figure 5.**
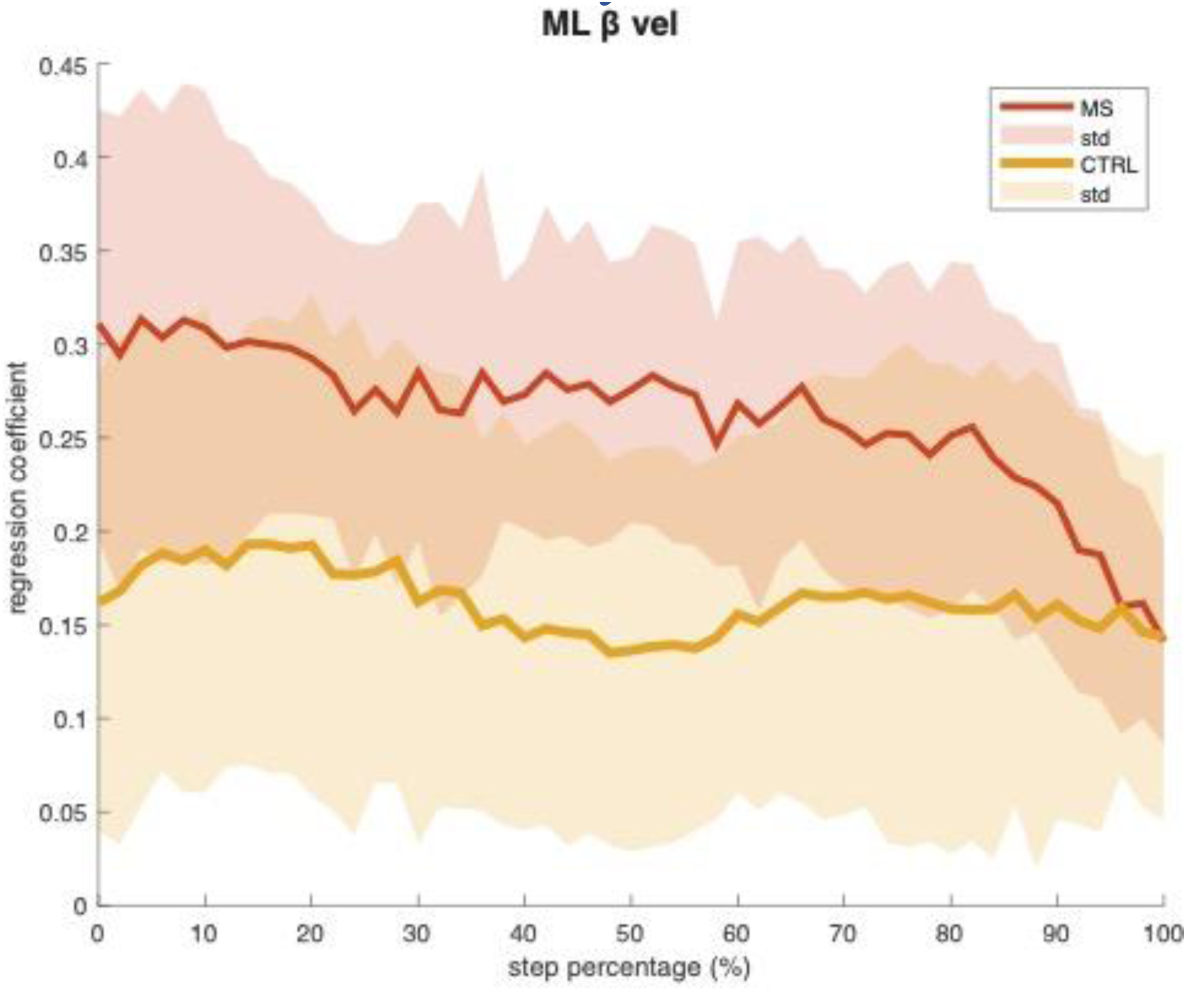
Mediolateral regression coefficient CoM_vel_. CoM_vel_ regression coefficient, as a measure of the strength of foot placement responses, across the step cycle, in PwMS (red) and controls (yellow).

**Supplementary Figure 6.**
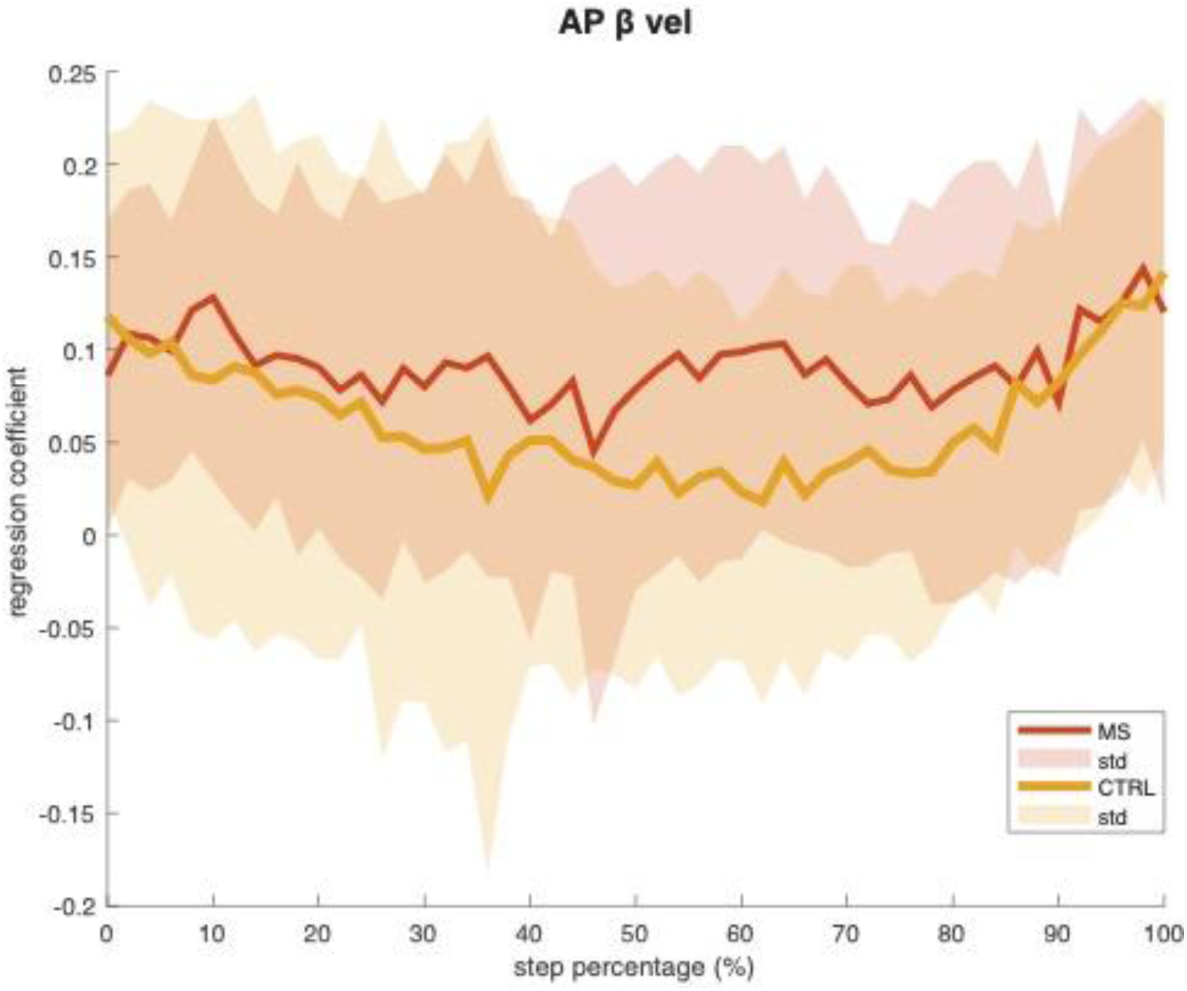
Anteroposterior CoM_vel_. CoM_vel_ regression coefficient, as a measure of the strength of foot placement responses, across the step cycle, in PwMS (red) and controls (yellow).

### Partial correlation CoM position

**Supplementary Figure 7.**
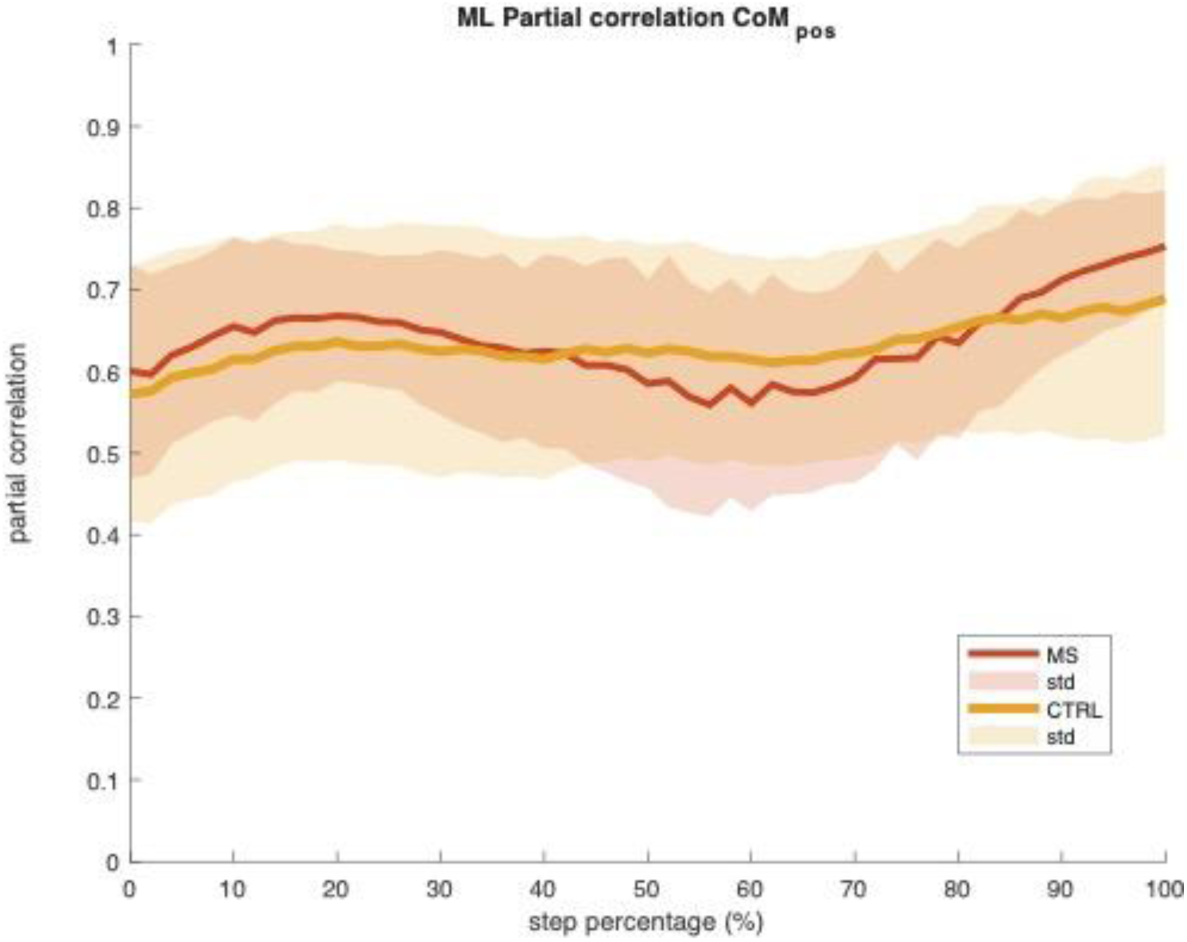
Mediolateral partial correlation of CoM_pos_. Partial correlation of CoM_pos,_, as a measure of feedback/control contribution across the step cycle in PwMS (red) and controls (yellow).

**Supplementary Figure 8.**
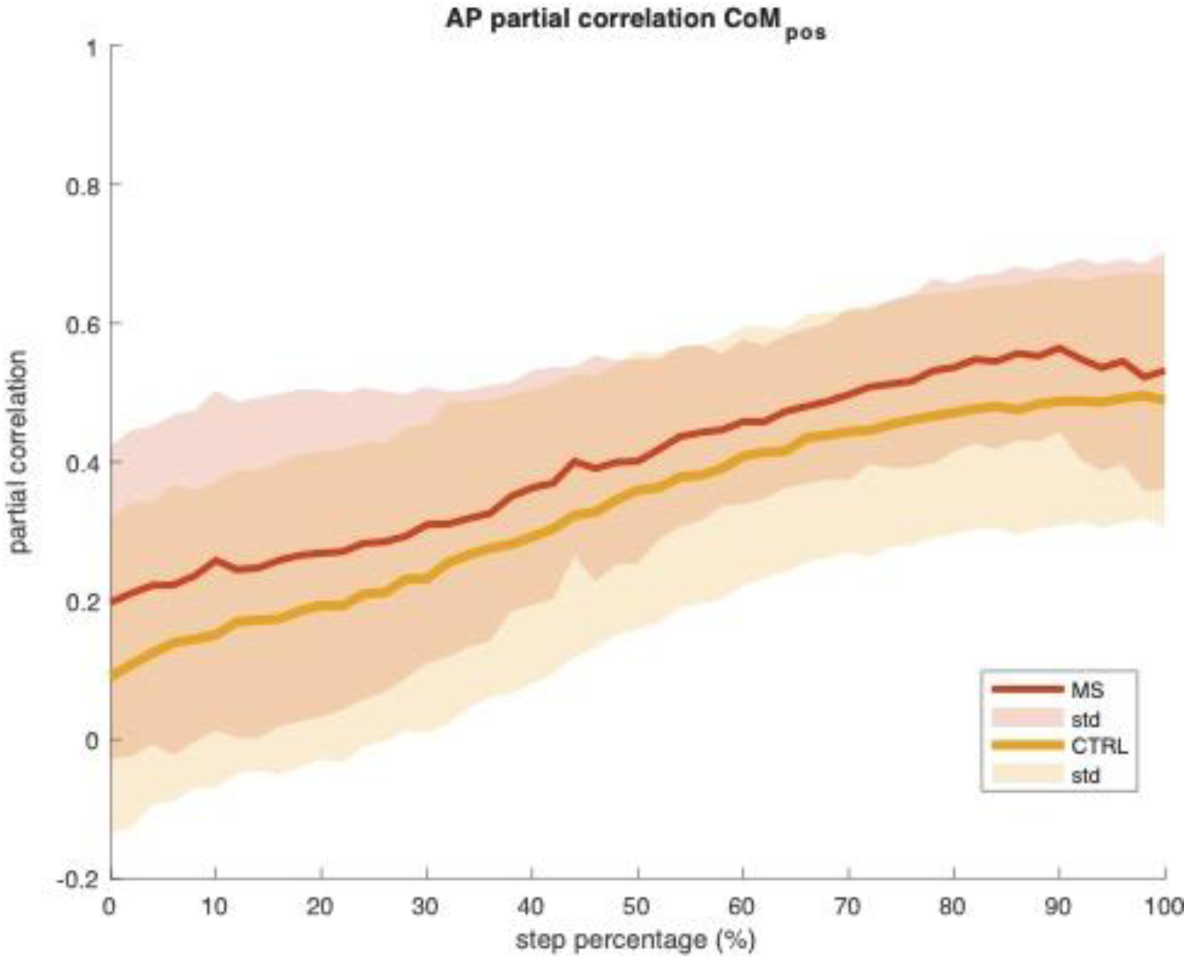
Anteroposterior partial correlation of CoM_pos_. Partial correlation of CoM_pos,_, as a measure of feedback/control contribution across the step cycle in PwMS (red) and controls (yellow).

### Partial correlation CoM velocity

**Supplementary Figure 9.**
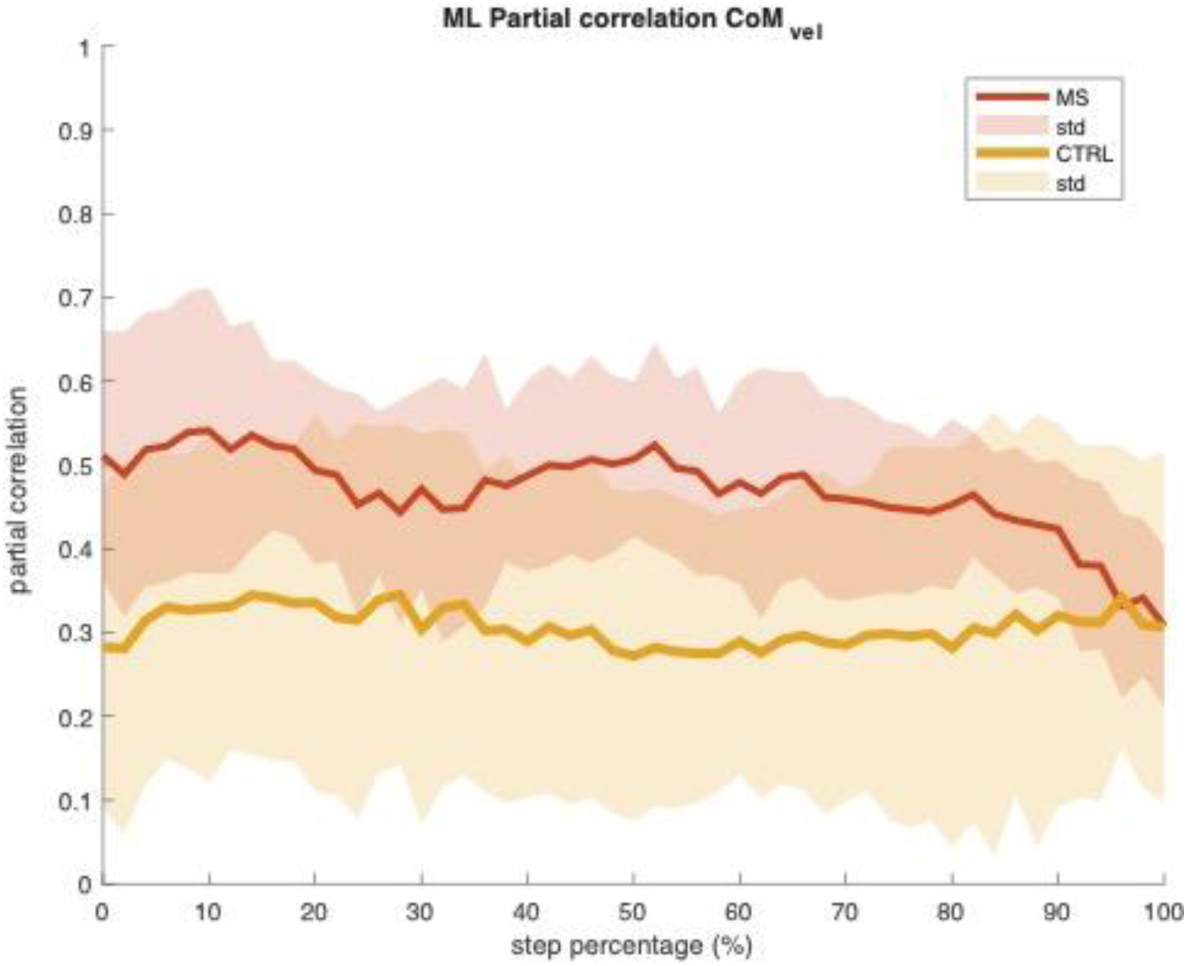
Mediolateral partial correlation of CoM_vel_. Partial correlation of CoM_vel_, as a measure of feedback/control contribution across the step cycle. The partial correlation is larger in PwMS (red) as compared to controls (yellow) during the feedback phase, but not at the end of the step.

**Supplementary Figure 10.**
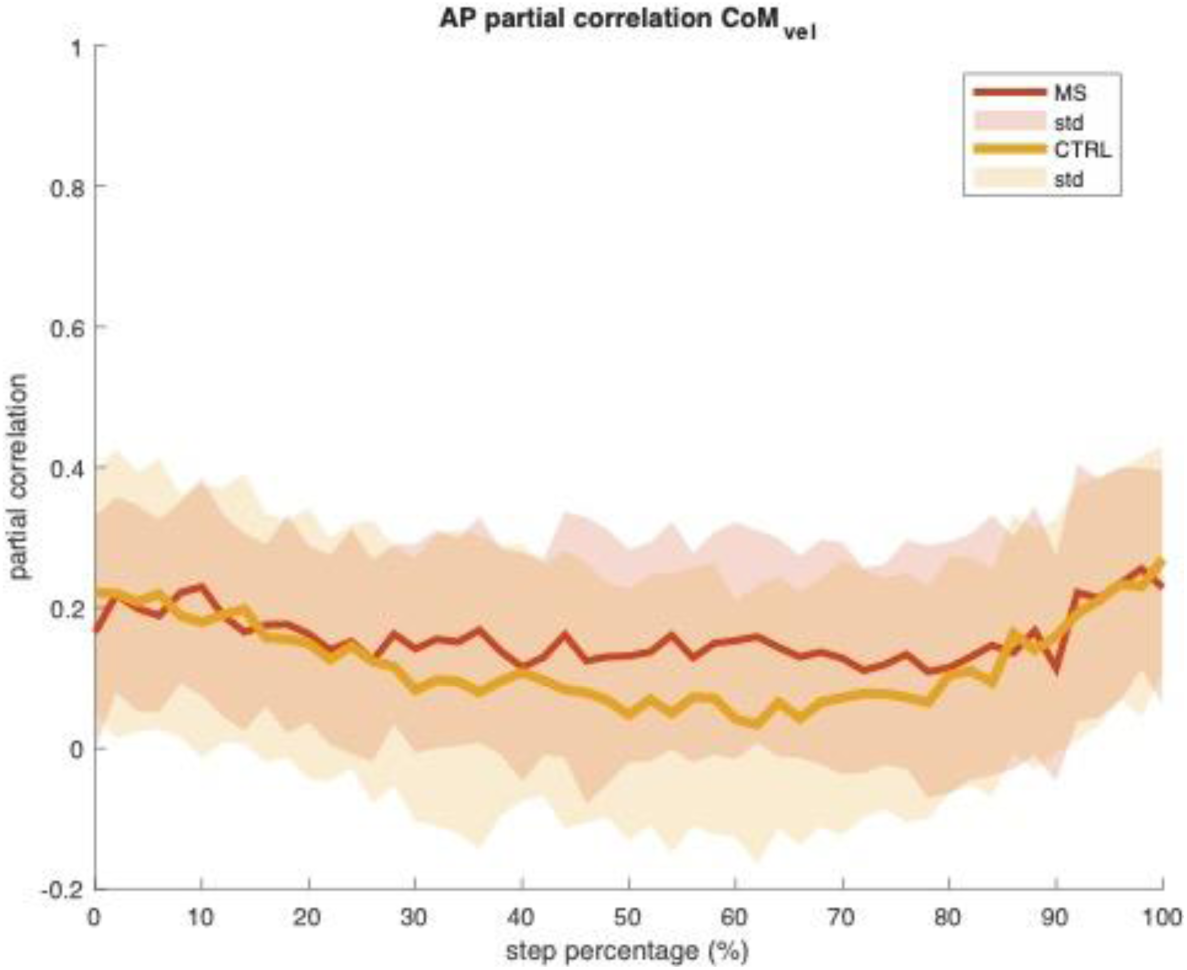
Anteroposterior partial correlation of CoM_vel_. Partial correlation of CoM_vel_, as a measure of feedback/control contribution across the step cycle, in PwMS (red) and controls (yellow).

